# Prevalence of Tuberculosis in Malnourished Children in Ethiopia: A Systematic Review and Meta-Analysis (2017-2025)

**DOI:** 10.64898/2026.09.21.26363606

**Authors:** Bihil Sherefedin, Nebiyou Yemanebrhane, Hassen Mamo

## Abstract

**Background:** Tuberculosis (TB), caused by *Mycobacterium tuberculosis* (MTB), remains a leading global public health challenge. Malnutrition is a major, modifiable driver of this vulnerability to the disease, creating a lethal, bidirectional link with TB. This s review aims to synthesize available evidence to estimate the pooled prevalence of TB and identify associated factors among malnourished children in Ethiopia, providing critical data to guide targeted, integrated public health interventions.

**Methods:** A systematic review and meta-analysis were conducted on studies reporting the prevalence of TB and associated factors among malnourished children in Ethiopia. Electronic databases including PubMed/Medline, EMBASE, CINAHL, ScienceDirect, Academic Search Complete, and Google Scholar were searched. In addition, bibliographies of relevant articles were reviewed, and a search for relevant conference abstracts was performed. Eligible studies were those published in peer-reviewed journals that specifically addressed TB prevalence among malnourished children. A random-effects model was employed to estimate the pooled prevalence of TB and its associated factors.

**Results:** A total of 746 studies were initially identified through database and catalog searches. Following screening and eligibility assessment, 12 studies were included in the final systematic review and meta-analysis. These studies, published between 2017 and 2024, collectively involved 5,749 participants. The smallest sample size (133) was reported in a study from the Oromia region, while the largest (1,201) was from the Amhara region. The pooled prevalence of TB among malnourished children was estimated at 16%. The highest prevalence (95%) was reported in a study conducted in Addis Ababa, while the lowest (6%) was from Harar. Considerable variation in prevalence was noted across the included studies.

**Conclusion:** This review resulted in the substantial prevalence of TB among malnourished children in Ethiopia, with a pooled prevalence of 16%. This finding highlights the urgent need for integrated healthcare interventions targeting both TB and malnutrition

## Introduction

Tuberculosis (TB) remains a leading global public health challenge, disproportionately affecting vulnerable populations, including children. In 2024, an estimated 10.7 million people developed TB worldwide, resulting in 1.23 million deaths, with children under 15 years accounting for approximately 11% of incident cases [1]. Despite this substantial burden, pediatric TB is critically under diagnosed; in 2024, only 64% of notified pulmonary TB cases in children were bacteriologically confirmed, reflecting persistent challenges in specimen collection, diagnostic access, and clinical recognition [1, 2]. Without timely treatment, mortality rates among children with TB can exceed 20–40%, particularly in infants and those with disseminated forms of the disease, such as TB meningitis [2].

Malnutrition is a major, modifiable driver of this vulnerability, creating a lethal, bidirectional link with TB. Under-nutrition manifesting as wasting, stunting, or underweight impairs cell-mediated immunity, reduces T-cell function, and disrupts the cytokine signaling necessary for MTB containment [3, 4]. Consequently, severely wasted children face a significantly elevated risk of developing active TB, with meta-analyses indicating up to a 3.5-fold increased odds compared to well-nourished peers [5]. Conversely, active TB aggravates nutritional depletion through reduced appetite, malabsorption, and increased catabolism [6].

The global co-burden of these conditions is stark. In 2024, an estimated 149 million children under five were stunted and 45 million were wasted worldwide, with the vast majority residing in sub-Saharan Africa and South Asia the same regions bearing the highest TB burdens [5, 7]. Despite the World Health Organization’s (WHO’s) and national guidelines advocating for integrated care, the translation into practice remains poor. For instance, in 2023, only 32% of eligible child contacts globally received TB preventive treatment (TPT), with coverage lowest in high-burden countries experiencing generalized food insecurity [1].

Ethiopia exemplifies this syndemic at scale. As one of the 30 high TB-burden countries, Ethiopia reported an estimated 154,000 incident TB cases in 2024, yet an 18% detection gap persists [1, 8]. Children constitute roughly 11% of these incident cases, yet TPT coverage among child contacts remains sub-optimal [1]. Concurrently, the country faces a severe malnutrition crisis; recent national data indicate that 33% of under five children are stunted, 7% are wasted, and 22% are underweight [8]. This overlap is most acute in drought- and, conflict-affected, and pastoralist regions (e.g., Somali, Afar, and Southern Ethiopia administrative Regions), where food insecurity, limited healthcare access, and high TB transmission converge [9]. Clinical evidence from Ethiopia confirms that malnourished children experience a 2-to 4-fold higher risk of TB progression, delayed sputum conversion, and increased mortality during treatment [10, 11]. A recent national survey further revealed that 23% of children admitted with severe acute malnutrition (SAM) had microbiologically confirmed or clinically diagnosed TB, a figure likely underestimated due to limited diagnostic capacity in stabilization centers [13].

Despite the 2023 UN High-Level Meeting on TB reaffirming the need for coordinated screening and combined interventions [9], and Ethiopia’s updated National TB Strategic Plan (2024–2028) prioritizing dual screening and routine nutritional support [10], routine TB screening remains largely absent in nutrition programs, and nutritional support is inconsistently integrated into pediatric TB care [14]. This fragmented approach undermines both TB control and child survival goals. Currently, evidence on the magnitude and determinants of TB among malnourished children in Ethiopia remains fragmented across disparate studies. Therefore, synthesizing the best available evidence is critical to inform policy, optimize resource allocation, and accelerate progress toward the ‘End TB’ and ‘Zero Hunger’ goals [5,9]. This systematic review and meta-analysis aims to provide up-to-date estimates of TB prevalence in this high-risk group, stratified by region, malnutrition severity, and diagnostic method, to guide targeted national and sub-national public health actions.

## Aim of the Study

To synthesize existing evidence on the burden and determinants of TB among malnourished children in Ethiopia through a systematic review and meta-analysis, in order to inform integrated public health strategies for dual screening, prevention, and management.

## Methods

### Search strategy

The search strategy, study selection, data extraction, and result reporting were conducted in accordance with the Preferred Reporting Items for Systematic Reviews and Meta-Analyses (PRISMA) guidelines [11, 12]. The PICO principle was adapted to guide the selection of search terms. Research articles included in this systematic review and meta-analysis were identified through searches in databases and repositories such as Google Scholar, Medline/PubMed, Cochrane Library, Web of Science, Hinari, ScienceDirect, African Journals Online, and university repositories (University of Gondar, Addis Ababa, Jimma, Hawassa, and Haramaya University). Search strategies were developed using Boolean operators (OR, AND, NOT) in combination with terms such as “prevalence,” “magnitude,” “proportion,” “burden,” “undernutrition,” “malnutrition,” “malnourishment,” “underweight,” “tuberculosis,” “children,” “child,” “adults,” and “Ethiopia.” Identified research articles were screened to ensure inclusion of all relevant literature. References were managed using Zotero software to organize citations and support the review process [13].

### Eligibility criteria

All studies conducted on TB and associated factors among malnourished children in Ethiopia were considered eligible. Articles published in the form of journal articles, master’s theses, and dissertations written in English were included. Only studies conducted within Ethiopia were considered.

### Operational Definitions (Source(s))

#### Children

Individuals below the age of 15 years.

#### TB

Defined as the presence of at least one sign or symptom suggestive of TB, including a persistent (> two weeks), non-remitting cough; weight loss or failure to thrive; persistent (> one week), unexplained fever (>38°C) recorded at least once; or persistent, unexplained lethargy or reduced playfulness. Microbiologically confirmed TB was defined by at least one positive culture with MTB speciation from a specimen representative of intrathoracic disease. Additional criteria included a history of prolonged fever and cough, failure to gain weight, loss of appetite, weight decline, and symptoms of extrapulmonary TB such as lymphadenopathy, seizures, abdominal pain, and contact with a confirmed TB case. Confirmed physician diagnoses based on patient chart reviews were also accepted.

#### Malnutrition

Refers to deficiencies or excesses in nutrient intake, imbalances in essential nutrients, or impaired nutrient utilization. It includes both undernutrition and overweight/obesity, along with diet-related non-communicable diseases.

#### Undernutrition

Manifests in four major forms: wasting, stunting, underweight, and micronutrient deficiencies.

### Outcome Measurement

The primary outcome was the prevalence of TB among children with malnutrition, calculated by dividing the number of children diagnosed with TB by the total number of participants included in the final analysis from the primary studies.

### Data collection and quality assessment

To assess the quality of the included studies, the Joanna Briggs Institute (JBI) quality appraisal checklist for observational studies was utilized [14]. A standardized data extraction checklist in Microsoft Excel was employed to extract relevant information. Zotero reference management software was used to consolidate search results from various databases and to eliminate duplicate entries. Research articles were initially screened based on their titles and abstracts. The full-text versions of the remaining articles were then reviewed for inclusion. Eligibility of the primary studies was determined using predefined inclusion and exclusion criteria. For the primary outcome prevalence of TB among children with undernutrition the data extraction checklist captured information such as the authors’ names, year of publication, study region, study design, sample size, response rate, and the number of TB cases identified. Data were also extracted for the secondary outcome concerning factors associated with undernutrition.

### Data analysis

The necessary information from each original study was retrieved using a format developed in Microsoft Excel. The data were subsequently exported to R Statistical Software (v4.1.2; R Core Team 2021) for analysis to determine the pooled effect size with a 95% confidence interval (CI). The Cochran Q test (Chi-squared statistic) and I^2^ statistic presented on forest plots were calculated to assess heterogeneity among the included studies. A p-value < 0.05 was considered statistically significant for Cochran’s Q test. I^2^ statistics, which range from 0% to 100%, were interpreted as indicating no (0%), low (25%), moderate (50%), or high (75%) levels of heterogeneity [13, 14]. A funnel plot and Egger’s test were also employed to assess potential publication bias.

### Ethics Statement

This study is a systematic review and meta-analysis of published data. No new data were collected from human participants, and therefore ethical approval was not required.

## Results

### Data Synthesis

A total of 746 studies were initially identified from various electronic databases and library catalogs. Among these, 405 records were excluded due to duplication. Screening of titles and abstracts led to the exclusion of an additional 318 articles that did not meet the inclusion criteria. Following full-text assessment of the remaining articles, 11 studies were excluded due to the absence of relevant outcome data. Ultimately, 12 studies met the eligibility criteria and were included in the final systematic review and meta-analysis (Fig. 1)

**Figure 1.**
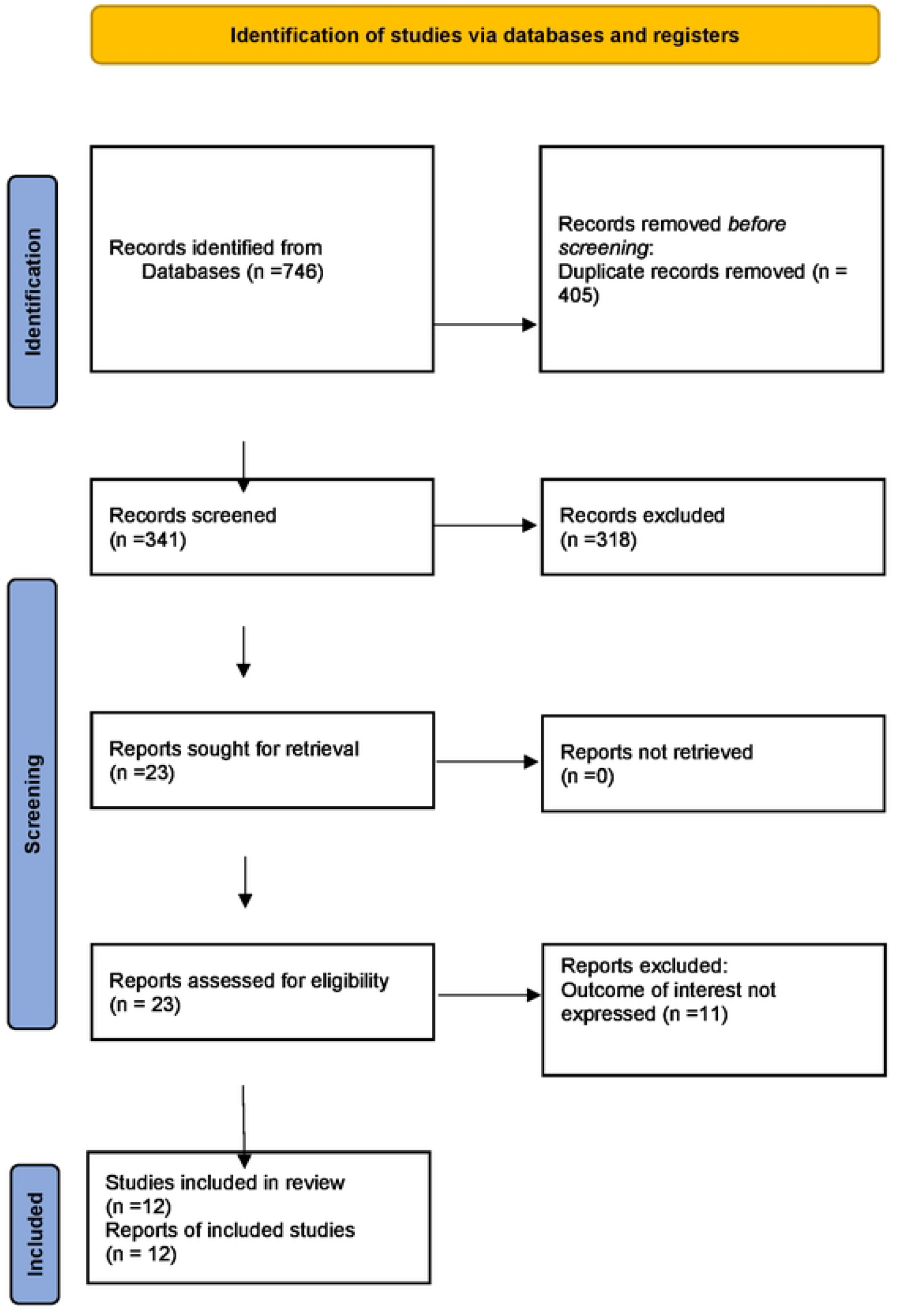
PRISMA flow diagram of included studies in the systematic review and meta analysis of the prevalence TB and associated factors of among malnourished children in Ethiopia.

#### Characteristics of included studies

All the 12 studies included in this systematic review and meta-analysis were published between 2017 and 2024 in peer-reviewed journals [15–26]. A total of 5,749 participants were encompassed across these studies. The smallest sample size was 133, reported in a study conducted in the Oromia [9], while the largest sample size was 1,201, reported in a study from the Amhara region [121]. Of the included studies, six employed a cross-sectional design [15,18,19,24–26], and six were retrospective cohort studies [17,20–23]. In terms of geographic distribution, four studies were conducted in the Amhara region [15,20,21,26], one in the former Southern Nations, Nationalities, and Peoples’ Region (SNNPR) [16], one in Oromia [16], two in Dire Dawa [18,27], in Harari [17,19], and in Addis Ababa [22,24] each (Table 1). Table 1 summarizes the key characteristics of these studies.

**Table 1.**
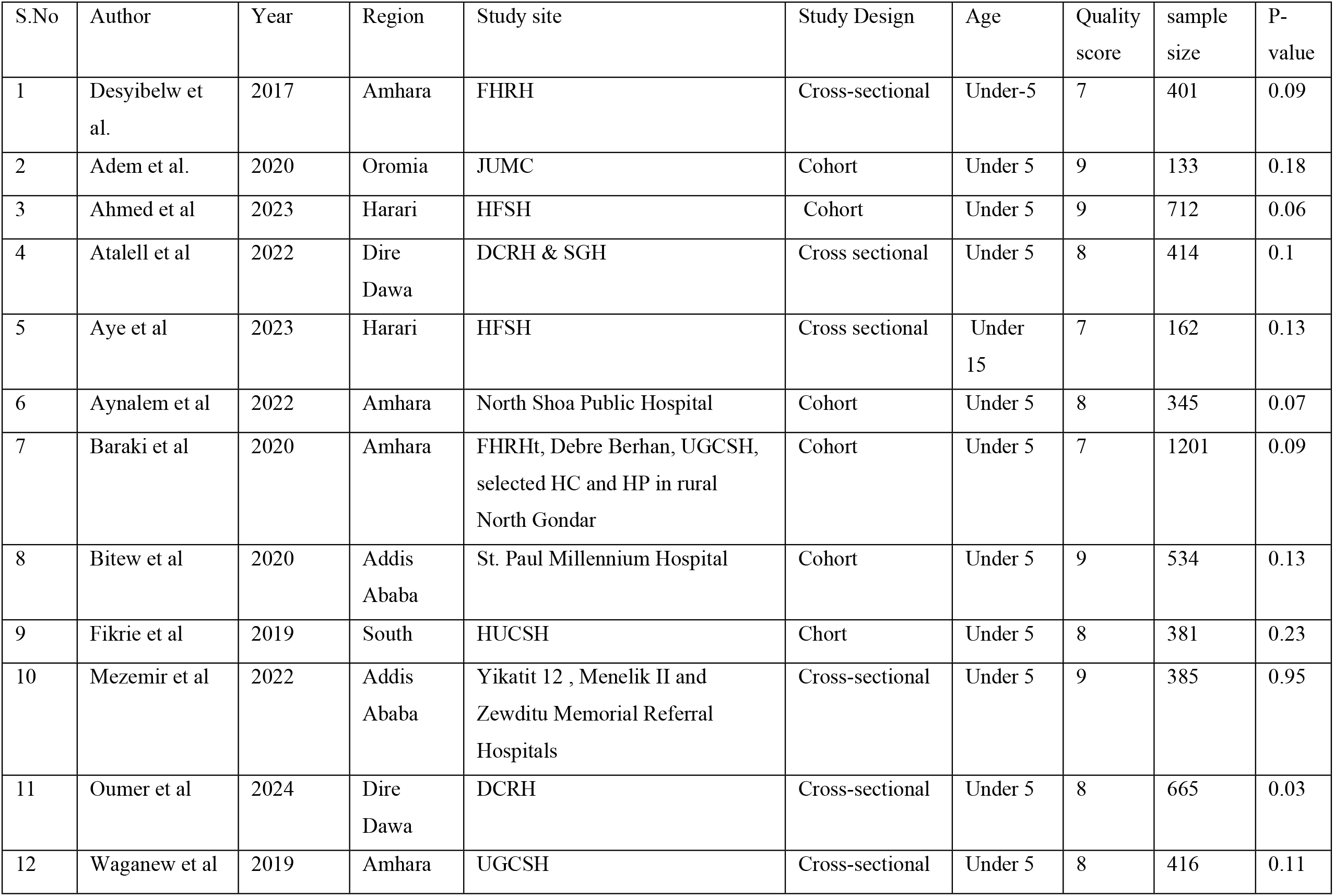
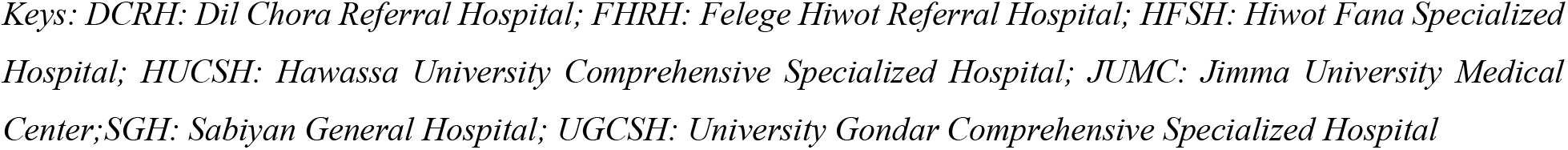
Characteristics of the included studies.

| S.No | Author | Year | Region | Study site | Study Design | Age | Quality score | sample size | P-value |
| --- | --- | --- | --- | --- | --- | --- | --- | --- | --- |
| 1 | Desyibelw et al. | 2017 | Amhara | FHRH | Cross-sectional | Under-5 | 7 | 401 | 0.09 |
| 2 | Adem et al. | 2020 | Oromia | JUMC | Cohort | Under 5 | 9 | 133 | 0.18 |
| 3 | Ahmed et al | 2023 | Harari | HFSH | Cohort | Under 5 | 9 | 712 | 0.06 |
| 4 | Atalell et al | 2022 | Dire Dawa | DCRH & SGH | Cross sectional | Under 5 | 8 | 414 | 0.1 |
| 5 | Aye et al | 2023 | Harari | HFSH | Cross sectional | Under 15 | 7 | 162 | 0.13 |
| 6 | Aynalem et al | 2022 | Amhara | North Shoa Public Hospital | Cohort | Under 5 | 8 | 345 | 0.07 |
| 7 | Baraki et al | 2020 | Amhara | FHRHt, Debre Berhan, UGCSH, selected HC and HP in rural North Gondar | Cohort | Under 5 | 7 | 1201 | 0.09 |
| 8 | Bitew et al | 2020 | Addis Ababa | St. Paul Millennium Hospital | Cohort | Under 5 | 9 | 534 | 0.13 |
| 9 | Fikrie et al | 2019 | South | HUCSH | Chort | Under 5 | 8 | 381 | 0.23 |
| 10 | Mezemir et al | 2022 | Addis Ababa | Yikatit 12 , Menelik II and Zewditu Memorial Referral Hospitals | Cross-sectional | Under 5 | 9 | 385 | 0.95 |
| 11 | Oumer et al | 2024 | Dire Dawa | DCRH | Cross-sectional | Under 5 | 8 | 665 | 0.03 |
| 12 | Waganew et al | 2019 | Amhara | UGCSH | Cross-sectional | Under 5 | 8 | 416 | 0.11 |

*Keys: DCRH: Dil Chora Referral Hospital; FHRH: Felege Hiwot Referral Hospital; HFSH: Hiwot Fana Specialized Hospital; HUCSH: Hawassa University Comprehensive Specialized Hospital; JUMC: Jimma University Medical Center; SGH: Sabiyan General Hospital; UGCSH: University Gondar Comprehensive Specialized Hospital*

#### Prevalence of TB among malnourished children in Ethiopia

The highest prevalence (P=0.95) was observed in a study conducted in Addis Ababa [22,24] and the lowest prevalence (P=0.03) was observed in a study conducted in Dire Dawa [10,12] (Fig. 2). The overall pooled prevalence, combining all studies, is 0.16 (95% CI: 0.15, 0.16). The I^2^ value of **99.8%** suggests substantial heterogeneity among the studies, indicating that they may have methodological or population differences. the p-value of <0.001 confirms statistical significance.

**Figure 2.**
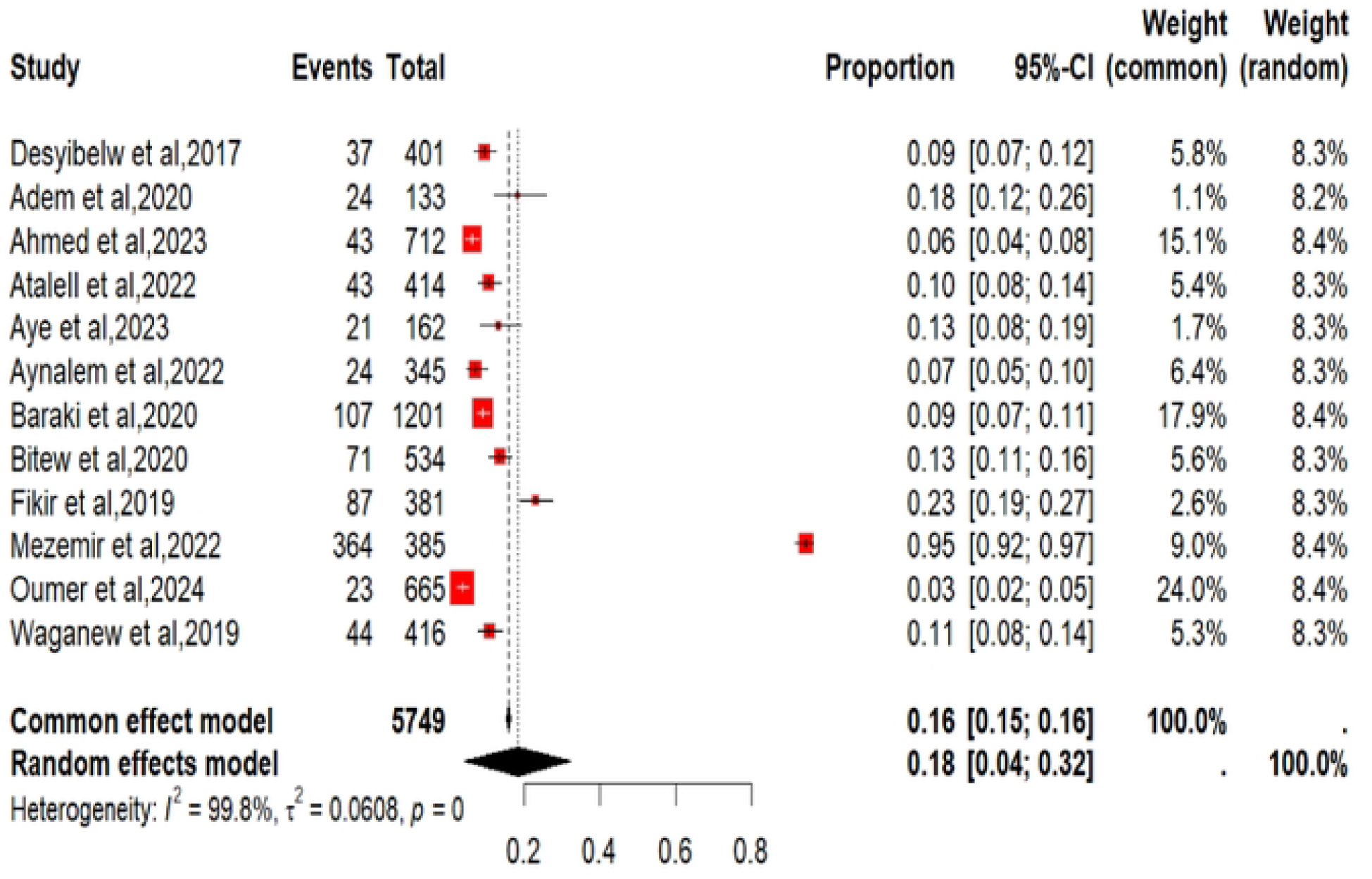
Forest plot of the pooled estimate of the prevalence of TB among malnourished children in Ethiopia.

#### Assessing Publication Bias in Meta-Analysis

The funnel plot shows asymmetry, there could be potential bias (Fig. 3). However, egger’s test indicated that P value of 0.90.

**Figure 3.**
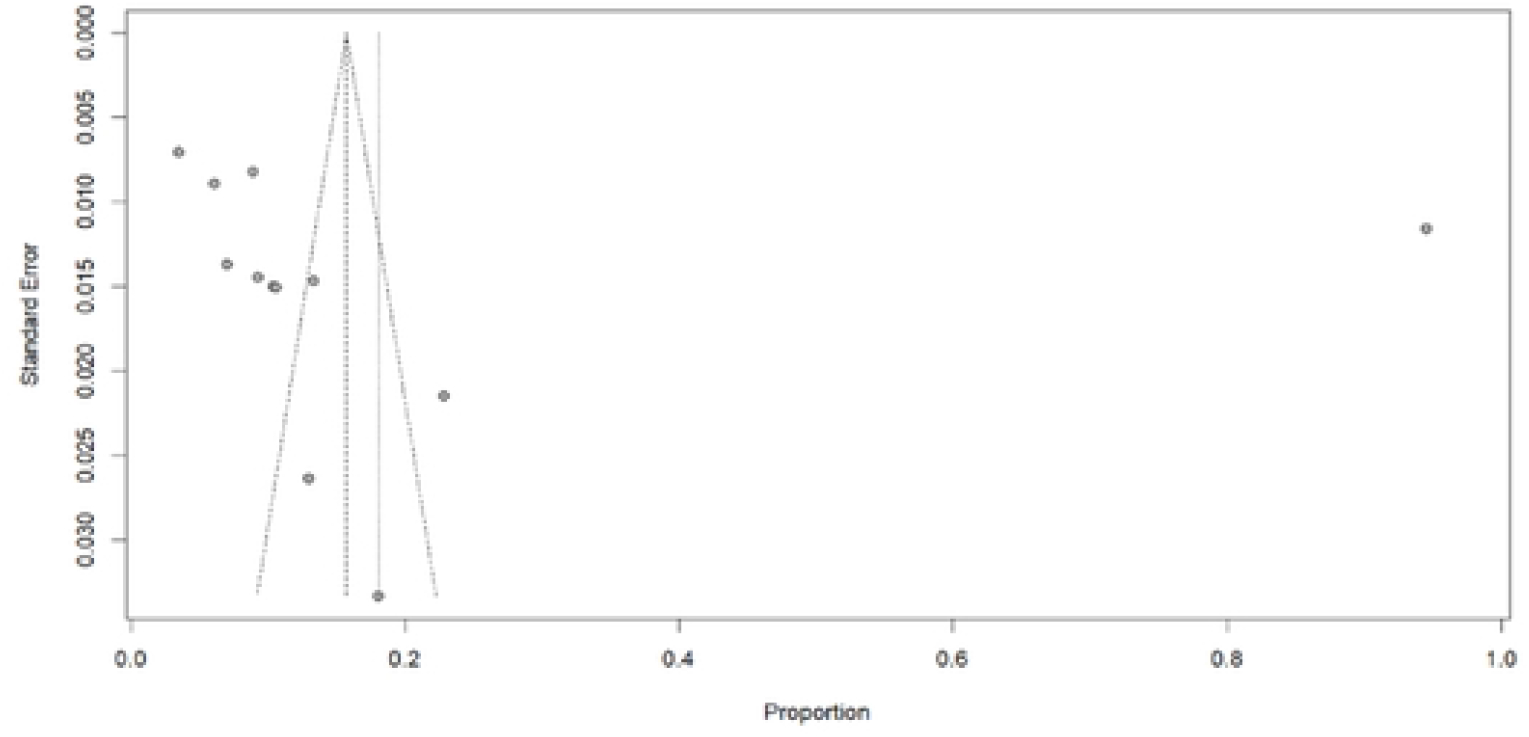
Funnel Plot Assessing Publication Bias in MetaAnalysis

## Discussion

This systematic review and meta-analysis was conducted to estimate the pooled prevalence of tuberculosis (TB) among malnourished children in Ethiopia and to identify associated factors. Twelve eligible studies were included, spanning multiple administrative regions including Addis Ababa, Amhara, Oromia, the former SNNPR, Dire Dawa, and Harari, thereby ensuring geographical diversity. The pooled prevalence of was estimated at 16% (0.16), indicating a substantial co-burden of TB and malnutrition in this vulnerable population. This finding is consistent with global evidence [3,11] demonstrating a strong bidirectional relationship between malnutrition and increased susceptibility to infectious diseases, including TB.

The observed prevalence estimates varied considerably across the included studies, ranging from 6% to 95%, reflecting substantial heterogeneity in the populations, settings, and methodologies examined. The highest prevalence (95%) was reported in Addis Ababa, whereas the lowest (6%) was observed in Harari. These disparities may be attributed to several factors, including differences in study design, sample size, regional socioeconomic conditions, healthcare infrastructure, and the baseline nutritional status of the enrolled children. The elevated prevalence in Addis Ababa may partly reflect urban-related determinants such as high population density, increased mobility, and comparatively accessibility different diagnostics, which collectively enhance case detection. Conversely, the lower prevalence reported in Harari may indicate under diagnosis, limited healthcare access, or constraints in diagnostic sensitivity rather than a true absence of disease.

The methodological diversity among the included studies further contributes to the observed variability. Six studies employed a cross-sectional design, measuring point prevalence of TB at the time of admission for severe acute malnutrition (SAM). For example, the cross-sectional study by Baraki et al. [12] (n = 1,201, Amhara region) yielded a moderate prevalence estimate of approximately 9%. The remaining six studies were retrospective cohort designs that tracked incident TB during follow-up periods. Notably, cohort studies conducted in Addis Ababa [23] captured infections potentially acquired during therapeutic feeding, highlighting critical gaps in infection prevention within overcrowded stabilization units. This distinction is methodologically significant: cross-sectional designs capture prevalent disease at a single time point, whereas cohort designs capture incident disease over time, including cases that may be missed in point-prevalence surveys. The inherent variability introduced by combining these two designs contributes substantially to the high statistical heterogeneity observed in this meta-analysis and underscores that TB risk in malnourished children is dynamic rather than static.

the meta-analysis showed significant heterogeneity (I^2^ = 99.8%), indicating that the observed differences in prevalence estimates were primarily attributable to true between-study variation rather than chance alone. While such heterogeneity complicates the interpretation of a single pooled estimate, it simultaneously underscores the necessity of examining regional, methodological, and contextual modifiers more closely. Despite the high heterogeneity and visual asymmetry observed in the funnel plot, Egger’s test yielded a non-significant p-value (p > 0.05), suggesting no strong statistical evidence of publication bias or small-study effects. Nevertheless, given the limited number of included studies and the considerable heterogeneity, further exploration through subgroup analysis or meta-regression is warranted to identify specific sources of variation.

From a public health perspective, a 16% TB prevalence among malnourished children constitutes a serious epidemiological concern. Given that undiagnosed pediatric TB carries a mortality risk exceeding 40% [2], and that malnourished children face a two-to four-fold increased risk of TB-related death [10], the present estimate implies that of Ethiopia’s approximately two million children under five years of age who experience SAM or moderate acute malnutrition (MAM) annually [8], more than 320,000 may be living with undetected TB. This undiagnosed reservoir sustains community transmission and contributes to Ethiopia’s estimated 18% TB case detection gap [1]. These findings reinforce the urgent need for integrated healthcare strategies that simultaneously address TB and malnutrition among vulnerable paediatric populations. Effective interventions should incorporate nutritional support alongside TB prevention, screening, and treatment, rather than addressing these conditions in isolation. Furthermore, the regional variability in TB prevalence calls for context-specific public health responses: regions such as Addis Ababa may benefit from enhanced targeted screening programmes leveraging existing diagnostic infrastructure, whereas areas like Harari may require investments in healthcare infrastructure and improvements in case detection to ensure accurate diagnosis and reporting.

Presently, however, service delivery remains fragmented. Nutrition programmes rarely incorporate routine TB screening [14], and nationally, only 32% of eligible child TB contacts receive TB preventive therapy (TPT) a proportion that is likely lower among children with SAM [1]. Conversely, TB programmes seldom assess nutritional status, despite national guidelines recommending nutritional support for all pediatric TB patients [16]. Furthermore, diagnostic capacity remains inadequate: access to GeneXpert is limited in rural primary hospitals [20], and child-friendly diagnostic tools such as stool Xpert Ultra and urine lipoarabinomannan (LF-LAM) assays remain scarce, forcing clinicians to rely on insensitive smear microscopy or non-specific clinical algorithms. These findings align with and support recent high-level policy commitments. The 2023 United Nations High-Level Meeting on TB prioritised coordinated screening for TB and undernutrition, particularly among children [15]. Similarly, Ethiopia’s 2024–2028 National TB Strategic Plan mandates dual screening of malnourished children for TB and HIV, as well as routine nutritional assessment for all pediatric TB patients [16]. The present evidence underscores the urgency of operationalizing these commitments at scale.

## Limitation

This review has several limitations that should be acknowledged. High statistical heterogeneity limits the precision of the pooled estimate and suggests caution in generalizing a single prevalence figure across all settings and methodological diversity between cross-sectional and cohort designs introduces variability.

## Conclusion

This systematic review and meta-analysis demonstrates a substantial burden of TB among malnourished children in Ethiopia, with a pooled prevalence of 16%. The finding highlights an urgent need for integrated TB nutrition screening, improved diagnostic capacity, and context-specific public health strategies across diverse regional settings. Addressing the intersection of TB and malnutrition in pediatric populations is essential to reducing mortality, improving case detection, and advancing Ethiopia’s progress toward the *End TB Strategy* targets. Future research employing larger sample sizes, standardized methodologies, and diverse diagnostic approaches is warranted to further clarify the factors contributing to this public health burden and to inform the development of effective, targeted interventions.

## Data Availability

All relevant data underlying the findings of this systematic review and meta-analysis are fully available without restriction within the manuscript and its Supporting Information files.

## Author Contributions

BS: Conceptualization, data extraction, statistical analysis, manuscript draft.

NY: Data curation, Software, Writing review & editing.

HM: supervision, critical review of the manuscript.

All authors have read and agreed to the published version of the manuscript.

## Data Availability Statement

All data generated or analyzed during this study are included in this published article.

## Acknowledgment

We would like to express our sincere gratitude to the Department of Microbial Sciences & Genetics and the School of Biomedicine & Laboratory Sciences at Addis Ababa University for providing a supportive academic environment for this work. We also extend our appreciation to Knocking Out TB by Community Visionaries of Ethiopia for their valuable insights and ongoing collaboration in pediatric TB research.

We are deeply grateful to the primary study authors whose published data made this systematic review and meta-analysis possible. Their dedication to generating evidence on the TB– malnutrition syndemic among Ethiopian children has been instrumental to this work.

Finally, we would like to thank Professor Hassen Mamo for his critical review, supervision, and valuable methodological guidance during the preparation of this manuscript, and Nebiyu Yemanebrhane for his contributions to data curation, statistical analysis, and manuscript editing.

